# Effectiveness of a digital clinical decision support algorithm for guiding antibiotic prescribing in pediatric outpatient care in Rwanda: A pragmatic cluster non-randomized controlled trial

**DOI:** 10.1101/2025.08.01.25332743

**Authors:** Alexandra V. Kulinkina, Victor P. Rwandarwacu, Joseph Habakurama, Ludovico Cobuccio, Martin Norris, Emmanuel Kalisa, Cassien Havugimana, Angelique Ingabire, Gillian A. Levine, Rainer Tan, Vincent Faivre, Alan Vonlanthen, Marie-Annick Le Pogam, Kaspar Wyss, Lisine Tuyisenge, Jean Claude S. Ngabonziza, Valérie D’Acremont

## Abstract

**Background:** Poor adherence to clinical guidelines and diagnostic uncertainty are key contributors to antibiotic overprescription, accelerating antimicrobial resistance—a major global health threat. We developed ePOCT+, a digital clinical decision support algorithm designed to assist primary care clinicians in the diagnostic process and therapeutic management of acutely ill children under 15 years of age. ePOCT+ integrates oxygen saturation and hemoglobin measurements to help detect severe illnesses, and the C-reactive protein rapid test to help distinguish between bacterial and viral infections. The goal of implementing ePOCT+ was to reduce antibiotic prescriptions without compromising clinical outcomes.

**Methods and findings:** We tested the effectiveness of ePOCT+ in a pragmatic, open-label, two-arm, parallel-group, cluster non-randomized controlled superiority trial in 32 Rwandan health centers. Sixteen sites implemented ePOCT+, while the remainder provided standard care, enabling the intervention-control comparison. After five months, the control group transitioned to the intervention, enabling a before–after comparison. The initial intervention group continued to use ePOCT+, enabling a longitudinal assessment. A total of 59,921 outpatient consultations were enrolled between 1 December 2021 and 30 April 2023; 47,822 new consultations were analyzed, after excluding referrals and re-attendance visits. The uptake of the intervention (percentage of registered cases managed using ePOCT+) was 75% on average.

In the per-protocol (PP) analysis, ePOCT+ use was associated with a significant reduction in antibiotic prescription rates from 70.5% to 24.5% in the intervention–control comparison (absolute difference −46.0, 95% confidence interval (CI) −52.5 to −39.5) and to 27.5% in the before–after comparison (−43.0, 95% CI: −52.5 to −39.5). The corresponding reductions were smaller in the intention-to-treat (ITT) analysis: −36.7 (95% CI: −42.1 to −31.3) in the intervention-control comparison and −26.7 (95% CI: −32.2 to −21.4) in the before-after comparison. Once reduced, prescription rates stayed low (25-40%) throughout the intervention period. Nonetheless, approximately 25% of prescribed antibiotics were not recommended by ePOCT+, indicating that with additional training and mentorship of clinicians, a bigger impact could be achieved on reducing unnecessary antibiotic prescriptions. Notably, only 6% of patients took additional medications not prescribed during the initial consultation, equivalent in both study arms.

Reduced antibiotic prescription rates did not increase the risk of clinical failure seven days after the initial consultation. In the PP analysis, adjusted relative risk [aRR] was 1.07, 95% CI 0.97–1.18 in the intervention-control comparison and 1.00, 0.92–1.10 in the before-after comparison. In the ITT analysis, aRRs were slightly elevated in both comparisons but remained within the predefined non-inferiority margin (upper 95% CI < 1.3): 1.14 (95% CI: 1.02–1.26) in the intervention-control comparison and 1.09 (95% CI: 1.02–1.18 in the before-after comparison.

**Conclusions:** In this pragmatic trial, the implementation of ePOCT+ substantially reduced antibiotic prescribing in pediatric outpatient care in Rwanda, without compromising clinical recovery. To reach meaningful scale, integrating into Rwanda’s national electronic medical record platform and ensuring that all clinical and reporting functions can be performed through a unified system, is a critical next step. Such integration could streamline service delivery, improve data quality, and promote more consistent, evidence-based care at scale.

**Trial registration:** Clinicaltrials.gov NCT05108831

## Introduction

Children in sub-Saharan Africa (SSA) continue to suffer from significant morbidity and mortality [1], with the majority of their medical needs being addressed at the primary care level [2].

Despite widespread adoption of the Integrated Management of Childhood Illness (IMCI) guidelines, their implementation has fallen short of expectations, largely due to poor adherence [3,4]. On average, only half of the recommended clinical assessments are performed during consultations [5]. Contributing factors include insufficient staffing [6,7], physical or cognitive overload and low motivation among healthcare workers [4], and limited access to essential diagnostic tools [8].

In the face of diagnostic uncertainty, time constraints, and decision fatigue, clinicians frequently resort to precautionary antibiotic prescribing [9]. Over 60% of pediatric outpatient consultations in SSA result in antibiotic use [10], often unnecessarily [11]. In addition to short-term side effects, altered microbiota, and compromised immunity [12], exposure to antibiotics in childhood can have long-term health consequences, such as allergies, obesity, and neurodevelopmental disorders [13]. Inappropriate antibiotic use also contributes to antimicrobial resistance (AMR) [14,15], a pressing global health threat now responsible for more deaths than malaria and HIV [16].

Digital clinical decision support algorithms (CDSAs) have been identified by the World Health Organization (WHO) as a strategy to strengthen adherence to evidence-based guidelines and improve the quality of care [17]. While CDSAs have shown promise in enhancing IMCI adherence [18–21], their impact on antibiotic prescribing remains inconsistent, with some studies showing a significant reduction [22–24], but not others [18,20]. Limitations of existing tools include restricted diagnostic scope [25], limited integration of point-of-care testing (e.g., for fever without source) [26], and poor uptake due to low digital literacy [27], lack of supervision, and inadequate clinical mentorship.

To address these gaps, we developed **ePOCT+** (electronic point-of-care tests plus), an advanced digital CDSA tailored to the Rwandan context [28]. Embedded in the **medAL-reader** tablet-based application [29], ePOCT+ supports the diagnosis and management of children across the full pediatric age range (1 day to 14 years) and a broad spectrum of acute conditions seen in outpatient primary care settings. The system integrates oxygen saturation and hemoglobin measurements (to help detect severe illnesses) and C-reactive protein rapid test (to help differentiate between viral and bacterial disease) into the diagnostic workflow and supports clinical mentorship via a real-time dashboard providing feedback on antibiotic prescription rates.

This study, which represents the main component of the <u>DYNAMIC project</u>, aimed to assess the effectiveness of ePOCT+ in reducing antibiotic prescribing for acutely ill children in Rwandan primary care settings. The project aligns with national priorities to strengthen pediatric care, promote rational antibiotic use [30,31], and advance the digital transformation of the health sector [32].

## Methods

### Study design and setting

The study was a pragmatic, open-label, two-arm, parallel-group, cluster non-randomized controlled superiority trial, conducted in 32 public health centers in Rusizi and Nyamasheke districts, Western Province, Rwanda. The trial employed a hybrid implementation design to support three distinct comparisons: (1) an intervention-control comparison; (2) a before-after comparison; and (3) a ten-month longitudinal assessment (Figure 1). The intervention-control comparison was the primary planned analysis; before-after comparison and longitudinal analysis are secondary. Health centers were grouped geographically into three implementation blocks (10 to 12 centers per block), with pragmatic allocation of half of the health centers within each block to the intervention (Group A) and half to the control (Group B) arm, enabling the intervention-control comparison. Allocation was stratified to ensure balance in terms of patient volume, infrastructure (e.g., electricity access), and use of a limited electronic medical record system in outpatient consultations. After five months, the intervention was extended to Group B, facilitating the before-after comparison. Group A continued using the intervention, enabling a longitudinal assessment (Figure 1).

**Figure 1:**
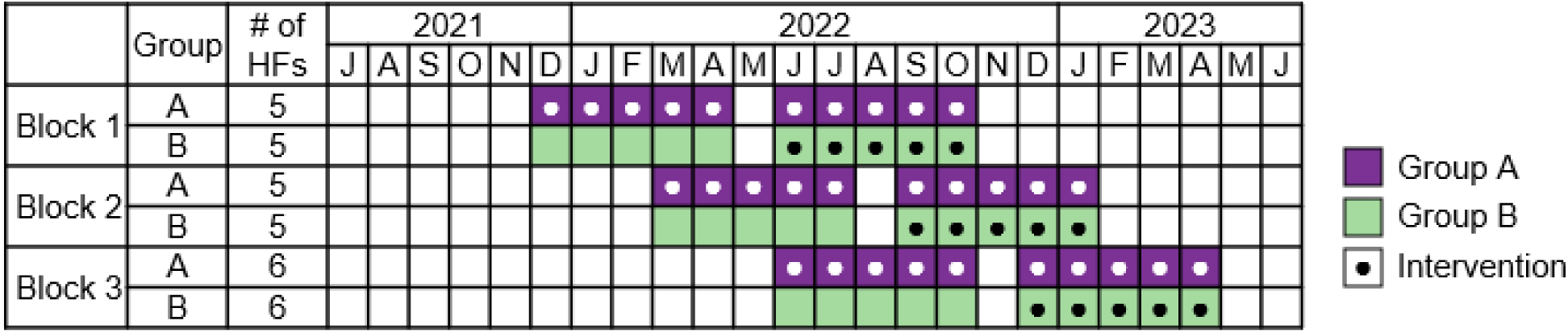
Implementation timeline. The figure shows staggered rollout of the intervention across the three blocks of participating health centers between 2021 and 2023. Group A received the intervention from the beginning (early and late intervention periods), while group B crossed over from control to intervention after five months. The three comparisons are: intervention-control (Group A vs. Group B, first five months), before-after (Group B pre-vs. post-intervention), and longitudinal (Group A early vs. late intervention).

The study area borders Lake Kivu and the Democratic Republic of Congo to the north-east and Nyungwe National Forest and Burundi to the south-west (Figure 2). This mountainous region experiences some of the highest precipitation rates in Rwanda [33]. The combined population of the two study districts is approximately 920,000, with 40% under the age of 15 [34].

**Figure 2:**
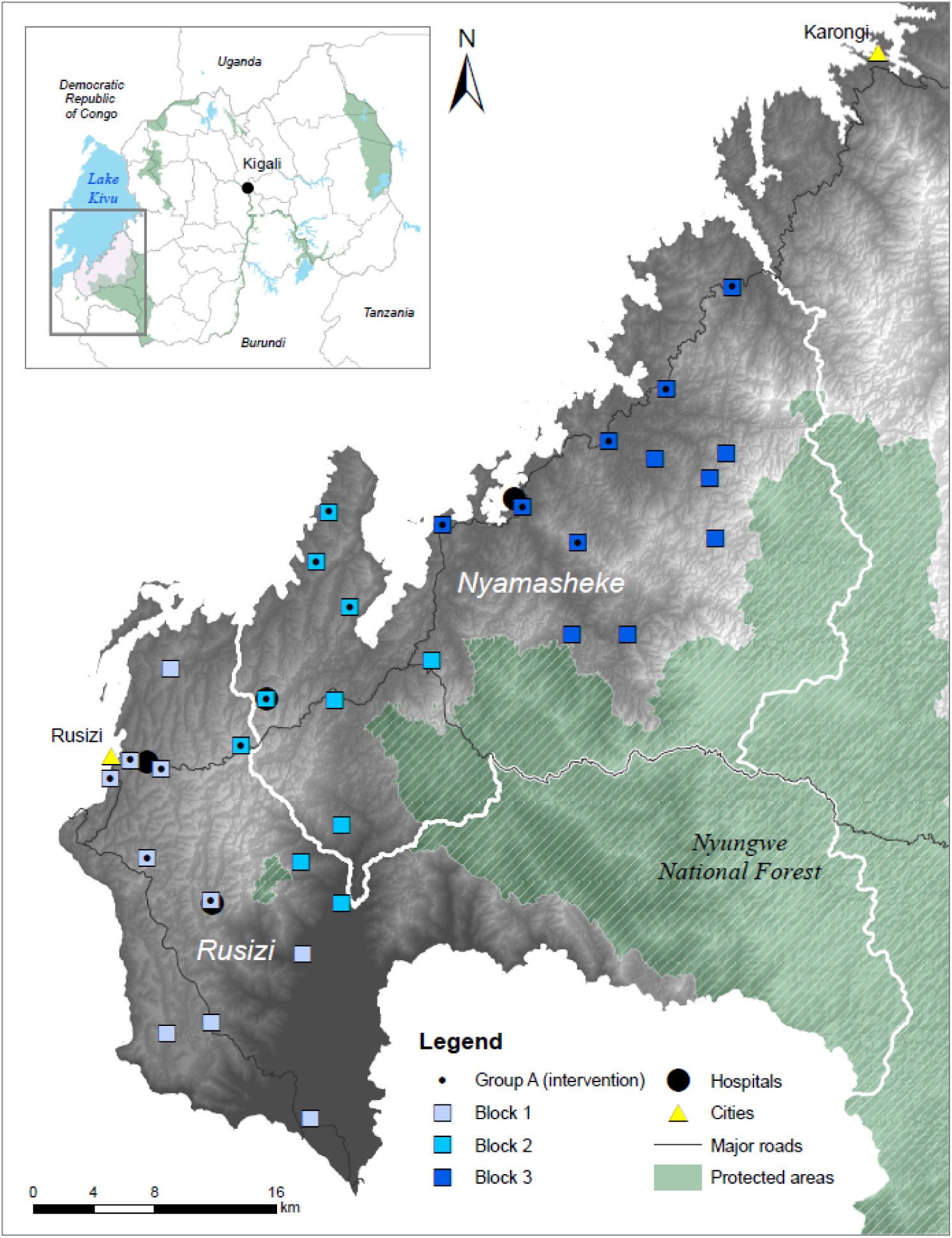
Map of the study area.

Nyamasheke district is more rural and remote, with 92% of the population residing in rural areas, as compared to Rusizi (67%) [34], which has substantial commercial and cross-border activity in and around Rusizi city. Rusizi has slightly higher prevalence of electricity coverage (67% vs. 62%), while mobile phone access is similar in both districts (around 80%) [34]. Both districts are highly endemic for malaria [35] and experience high prevalence of malnutrition and anemia, while HIV among children is rare, with a prevalence of 0.2% [36]. Up to 90% of the disease burden in Rwanda is treated at the primary care level, comprised of health centers, health posts, and community health workers (CHWs) [35]. Healthcare costs in the public sector are covered by health insurance, most often the government scheme Mutuelle de Sante, with high coverage (>98%) [34].

### Participants

Eligible facilities included public health centers, operated either by the government alone or in partnership with faith-based organizations. Health posts were excluded due to limited patient volumes and inconsistent staffing. Children aged 1 day to less than 15 years presenting with acute medical or surgical conditions were eligible for inclusion. Children presenting for routine preventive care or chronic disease follow-up, or those for whom informed consent could not be obtained, were excluded. In Rwandan health centers, pediatric consultations are conducted by nurses with A2, A1, or A0 qualifications [37] – these were the target users of the CDSA. Consultations for children under five are conducted in dedicated rooms, while older children are typically seen in general consultation areas together with adults.

### Sampling, randomization and masking

Out of 38 eligible health centers, 32 were selected, after excluding four that were difficult to reach and two that were involved in other research studies. Group allocation was performed geographically to limit contamination while maintaining balance. A non-randomized study design was chosen so that the intervention could be rolled out gradually to mimic how it would be done in government programs. Research assistants assessing clinical outcome at day 7 were blinded to the intervention allocation. Masking of clinicians, other study staff, and participants was not possible due to the nature of the intervention.

### Intervention

The ePOCT+ CDSA, implemented on tablets in the medAL-*reader* application [29], was integrated into the routine outpatient workflow in the participating health centers (Figure 3). In the intervention arm, clinicians used the algorithm to guide history taking, examination, and diagnostic decisions, including point-of-care testing not normally offered in the Rwandan primary care setting (hemoglobin, C-reactive protein, and pulse oximetry). All lab tests (routine and additional) were performed in accordance with the algorithm recommendations by the laboratory technician, while oxygen saturation was measured by the clinicians in the consultation room.

**Figure 3.**
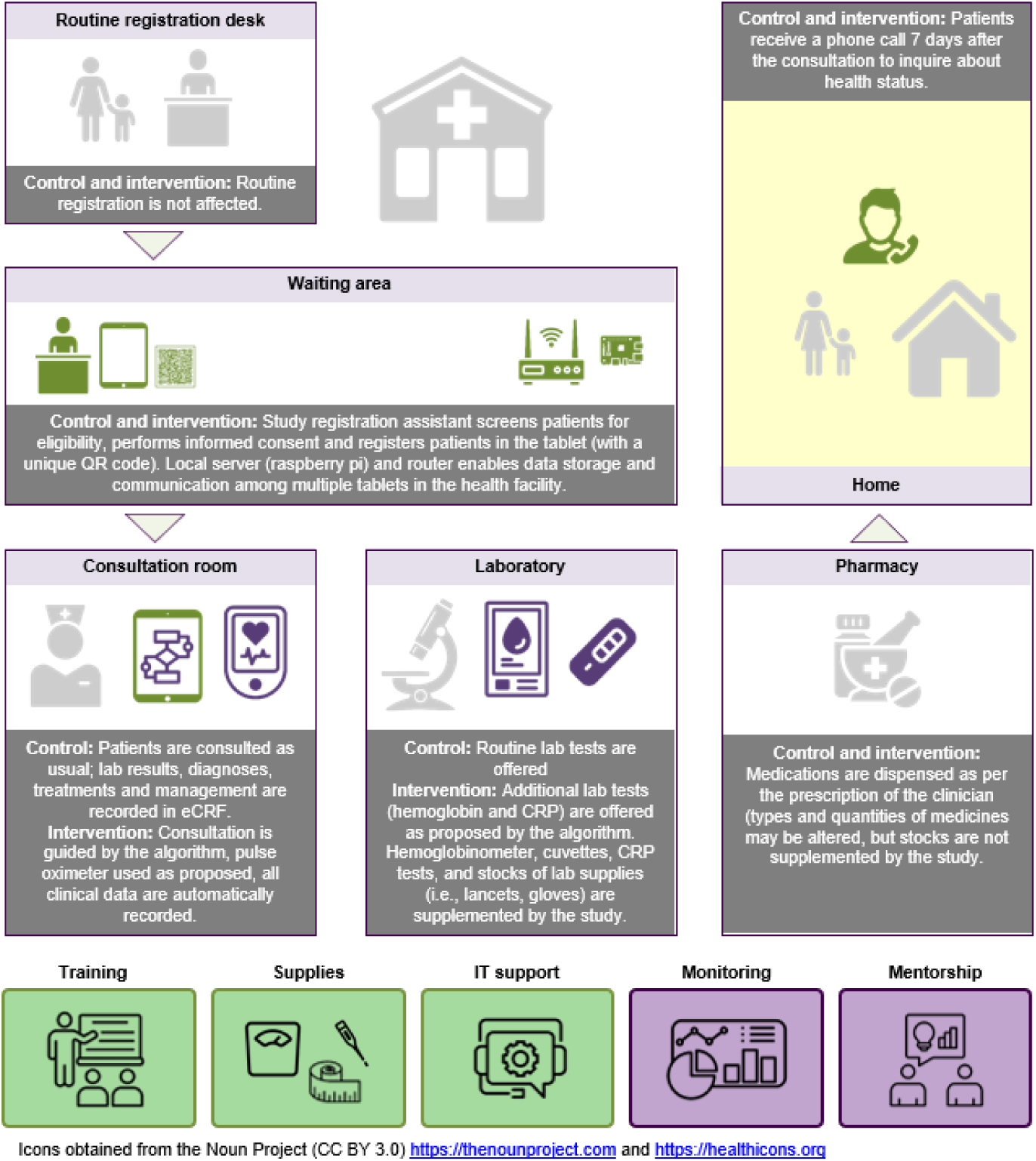
Clinical workflow in the intervention and control arms. The figure illustrates the clinical workflow for both study arms. In both arms, trained study registration assistants screened patients for eligibility, obtained informed consent, and registered patients using a tablet-based system. In the control arm, consultations followed routine clinical practice, and data were entered into eCRFs. In the intervention arm, consultations were guided by the ePOCT+ algorithm, which integrated clinical assessment with pulse oximetry and, additional point-of-care tests (hemoglobin and CRP), where indicated. Laboratory and pharmacy procedures remained similar across groups, though laboratory supplies were supplemented for the intervention. All participants received a follow-up call on day 7 to assess clinical status. Items in light gray are routine and are not affected by the intervention; items in green are provided to both intervention and control arms, items in purple are provided only to the intervention arm. eCRF – electronic case report form; CRP – C-reactive protein.

Based on the information entered in medAL-*reader*, the algorithm proposed diagnoses and corresponding treatment options aligned with national pediatric guidelines [28]. Clinicians could accept or override suggestions. In the control arm, routine care continued, and clinicians used the same medAL-*reader* application (without decision support) to enter lab test results, diagnoses, treatments and managements into an electronic case report form (eCRF).

Both arms received equivalent two-day clinical refresher training, but the intervention group received an additional one day of instruction on the CDSA. Supplies such as thermometers, weighing scales, and IT support were provided to all sites, with additional supplementation of lab consumables (e.g., gloves, lancets, alcohol swabs) in the intervention arm. A real-time monitoring dashboard provided data of intervention health facilities on key clinical indicators such as antibiotic prescribing, malaria management, and taking of anthropometric measurements. Data was presented in a way that allowed comparison of health facilities to each other (benchmarking). The implementation team used the dashboards to conduct clinical mentorship, which included reviewing the trends together with health facility clinical teams.

### Study procedures

### Eligibility screening and registration

Trained registration assistants co-hired by the study team and the health centers screened children under 16 years of age for eligibility: age below 15 years and presence of an acute illness. Those meeting the criteria and whose caregiver provided written informed consent were registered in the medAL-*reader* application, creating a medical case for the clinician in the consultation room to complete. Registration data included demographics and contact information for collection of outcomes. Aggregated screening summaries, including notes on study-specific issues, were submitted daily to the study team, who monitored data quality and provided feedback.

### Medical consultation (on day 0)

In the intervention arm, all clinical consultation data were captured through the real-time use of the algorithm. In the control arm, lab tests, diagnoses, and treatments were recorded in the eCRF. In addition, on the last page in the application, the clinicians in both arms reported whether they prescribed an oral or systemic antibiotic, and whether they referred the patient for inpatient admission (including overnight observation in the health center or referral to the hospital) or recommended a follow-up consultation. Filling this last page defined a completed case, or adherence to the intervention for per-protocol analysis.

### Follow-up phone call (on day 7)

Independent research assistants conducted telephone follow-up with caregivers on day 7 (range 6-14 days) to assess the clinical outcome and treatment seeking behavior since the initial consultation. Outcome information was recorded in standardized REDCap web forms by five callers; patients were randomly allocated to the callers who were blinded to the intervention status. In a minority of cases where caregivers were unreachable, CHWs were engaged to visit the household of the patient. However, data from these visits were excluded from primary analyses due to inconsistencies discovered during exploratory analyses (Figure S1, Supporting Information).

### Outcomes

The primary outcome was the proportion of children prescribed antibiotics at initial consultation (day 0). Secondary outcomes included referral or re-attendance visit recommendation at day 0 (as reported by clinicians in the application); clinical failure (defined as ‘not cured’ and ‘not improved’, or non-referred secondary hospitalization), hospitalization, severe outcomes (non-referred secondary hospitalization or death), taking of additional medications, and re-attendance by day 7 (as reported by caregivers); completion of referral or re-attendance visit; and malaria testing and treatment. Outcome definitions and relevant analysis populations are detailed in the Results and trial registry.

### Sample size

The sample size was calculated to detect a ≥25% relative reduction in antibiotic prescribing in the intervention-control comparison with 80% power and α = 0.05, assuming an ICC of 0.025, cluster size of 660 patients over a period of three months, and a conservative baseline antibiotic prescription rate estimate of 35%. A minimum of 12 centers per arm was required; all 32 eligible centers were enrolled to enhance geographic coverage and improve estimates of secondary outcomes. After extending the data collection period from three to five months due to lower-than-expected enrolment, the final per-protocol sample averaged 589 patients per cluster.

### Statistical analysis

Mixed-effects logistic regression models were used for all outcomes, with health center as a random effect. In models where singularity was observed, the random effect was removed without affecting estimates. Patient random effect was not accounted for due to relatively few patients having multiple consultations and resulting model convergence issues. Time random effect was also omitted, it did not affect estimates. All models were adjusted for age, sex, district, and five presenting complaint categories (fever, respiratory, gastrointestinal, ear nose throat and mouth, skin). Effect differences across these variables were also explored in sub-group analyses. Models for day 0 outcomes were also adjusted for monthly enrollment (quintile); day 7 models were adjusted for time to follow-up and caller identity. These additional covariates were not specified in the study protocol but were important in exploratory analyses (Figure S1, Supporting Information). Relative risks and absolute differences were estimated using marginal probabilities [38]; confidence intervals were derived via the Delta method [39].

Primary analyses were performed on the per-protocol (PP) population (cases completed in the application) for day 0 outcomes and on the complete cases (PP population with follow-up information) for day 7 outcomes. A conservative ITT analysis for antibiotic prescription assumed an antibiotic was prescribed when the CDSA was not used. Superiority in antibiotic prescription was defined as a 25% or more relative reduction between routine care and ePOCT+.

Noninferiority for clinical failure was defined as the upper limit of the 95% confidence interval (CI) of the adjusted risk ratio (aRR) <1.3. Prespecified subgroup analyses stratified the results by sex, age group, and presenting complaint. Due to all questions set mandatory in the data collection instruments, there was no missing data in the analysis variables. All analyses were conducted in R (version 4.2.2) [40] using the CRTSize package (version 1.2) [41] for sample size estimation and custom scripts for statistical modelling. The statistical analysis plan and study protocol can be found in the clinicaltrials.gov registry. De-identified data used in the analysis can be found at https://doi.org/10.5281/zenodo.15870974.

### Ethics and inclusion

The study was approved by the Rwanda National Ethics Committee (original protocol 752/RNEC/2020; amendment 246/RNEC/2023) and the Swiss Vaud Cantonal Ethics Commission (CER-VD 2020-02799). The trial was registered on ClinicalTrials.gov (NCT05108831; https://clinicaltrials.gov/study/NCT05108831). Additionally, the study was reviewed and approved by the National Health Research Committee (NHRC/2020/Prot/031) and the National Institute of Statistics of Rwanda (0654/2020/10/NISR). Subsequently, the Ministry of Local Government granted approval to work in Rusizi and Nyamasheke districts. Upon introduction of the project to the mayors’ offices and district hospitals, letters of collaboration were signed with each participating health center. Finally, the study was introduced to the Sector Council, and community engagement events were held in the catchment villages of each health center, with over 7,000 participants.

From the conceptualization stage, the study team worked with the Ministry of Health (MoH; via a Memorandum of Understanding) and the Rwanda Biomedical Center (RBC; via a collaboration agreement). A steering committee, with representatives from MoH, RBC, WHO, UNICEF, Rwanda Food and Drug Administration, Rwanda Pediatric Association, Rusizi and Nyamasheke districts, oversaw the study. Study approaches were regularly discussed with the national Maternal and Child Health Technical Working Group. At the district level, the team participated in quarterly meetings of the Joint Action Development Forum (JADF). Upon conclusion of the study, the findings were presented to the participating health centers, district hospitals, District Health Management Teams, relevant national programs at RBC and MoH and finally to a broader audience of stakeholders in a <u>dissemination event</u> in Kigali in March 2024.

## Results

### Baseline characteristics of health facilities and patients

A total of 32 health centers were included in the trial (Figure 4): 16 in the intervention group (Group A, ePOCT+) and 16 in the control group (Group B, routine care). The phased rollout (Figure 1) embedded ePOCT+ into the routine consultation workflow (Figure 3). Health centers were balanced in terms of district (Rusizi vs. Nyamasheke), ownership (public vs. faith-based partnership), and service readiness scores. Power supply and use of computers in the consultation rooms were also similar. Group A had slightly more outpatient staff (mean 9.3 vs. 7.8) and lower median monthly consultation volumes for children under five (236 vs. 286), resulting in marginally lower patient recruitment (Table 1).

**Figure 4.**
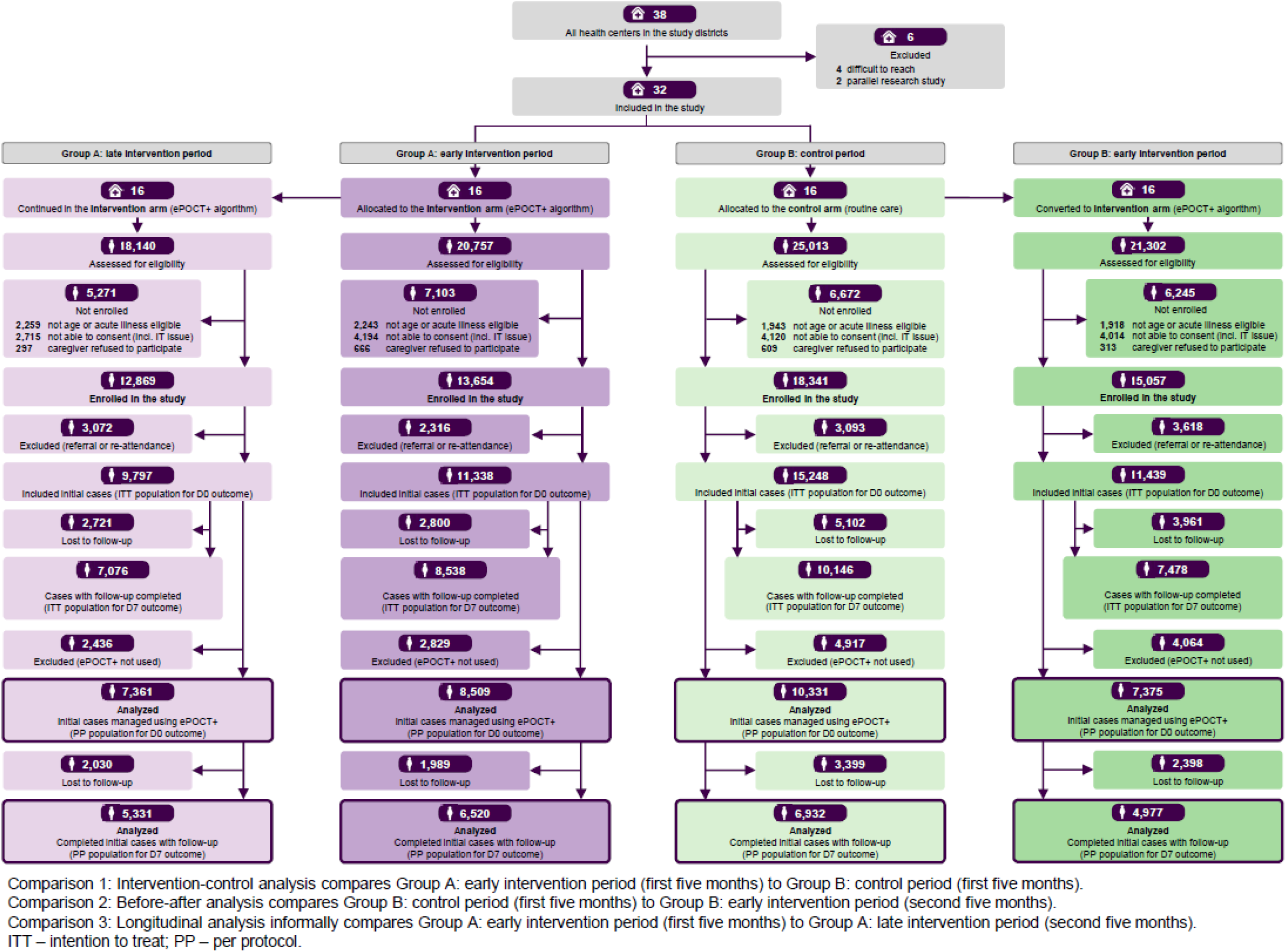
CONSORT flow diagram of participant inclusion by trial phase.

**Table 1.** Characteristics of participating health facilities and enrolled pediatric patients.

|  | A: early intervention<sup>a</sup><br>(ePOCT+) | A: late intervention<br>(ePOCT+) | B: control<sup>a,b</sup><br>(routine care) | B: early intervention<sup>b</sup><br>(ePOCT+) |
| --- | --- | --- | --- | --- |
| Health centers | n = 16 | n = 16 | n = 16 | n = 16 |
| District |  |  |  |  |
| Rusizi | 6 | 6 | 7 | 7 |
| Nyamasheke | 10 | 10 | 9 | 9 |
| Ownership |  |  |  |  |
| Public | 7 | 7 | 7 | 7 |
| Faith-based partnership | 9 | 9 | 9 | 9 |
| Service Availability and Readiness Assessment (SARA) score<sup>c</sup> |  |  |  |  |
| General services score, % (mean, SD) | 71.5 (7.0) | 71.5 (7.0) | 74.3 (6.7) | 74.3 (6.7) |
| Pediatric services score, % (mean, SD) | 74.7 (8.2) | 74.7 (8.2) | 76.1 (8.1) | 76.1 (8.1) |
| Power supply<sup>d</sup> |  |  |  |  |
| Uninterrupted | 9 | 9 | 7 | 7 |
| Outages <2 hrs per day | 6 | 6 | 9 | 9 |
| Outages ≥2 hrs per day | 1 | 1 | 0 | 0 |
| Using computer in consultation room for clinical data entry<sup>d</sup> | 4 | 4 | 4 | 4 |
| Staffing: # of OPD clinicians (mean, SD)<sup>d</sup> | 9.3 (2.4) | 9.3 (2.4) | 7.8 (2.3) | 7.8 (2.3) |
| Patient load: # of <5 patients per facility per month (median, IQR)<sup>e</sup> | 236 (159, 349) | 236 (159, 349) | 286 (225, 347) | 286 (225, 347) |
| Enrollment: # of <15 patients per facility per month (median, IQR) | 151 (124, 222) | 155 (117, 194) | 207 (130, 294) | 173 (125, 231) |
| Number of health facility days when the study was operating<sup>f</sup> | 1,639 | 1,639 | 1,639 | 1,639 |
| Patients enrolled | 1,515 | 1,508 | 1,541 | 1,541 |
| Patients not enrolled: registration assistant off duty | 43 | 54 | 46 | 37 |
| Patients not enrolled: no trained health worker to use the app | 67 | 62 | 37 | 38 |
| Patients not enrolled: power or IT issue | 11 | 15 | 10 | 17 |
| Patients not enrolled: another issue | 3 | 0 | 5 | 6 |
| **Patients (enrolled; intention-to-treat)** | **n = 13,654** | **n = 12,869** | **n = 18,341** | **n = 15,057** |
| Sex (n, %) |  |  |  |  |
| Female | 7,067 (51.8%) | 5,998 (46.6%) | 9,344 (50.9%) | 8,061 (53.5%) |
| Male | 6,587 (48.2%) | 6,871 (53.4%) | 8,997 (49.1%) | 6,996 (46.5%) |
| Age group (n, %) |  |  |  |  |
| <2 months | 611 (4.5%) | 535 (4.2%) | 946 (5.2%) | 699 (4.6%) |
| 2-59 months | 8,852 (64.8%) | 8,376 (65.1%) | 11,678 (63.7%) | 10,324 (68.6%) |
| 5-14 years | 4,191 (30.7%) | 3,958 (30.7%) | 5,717 (31.2%) | 4,034 (26.8%) |
| Consultation type (n, %) |  |  |  |  |
| New | 11,338 (83.0%) | 9,797 (76.1%) | 15,248 (83.1%) | 11,439 (76.0%) |
| Follow-up | 2,292 (16.8%) | 3,066 (23.8%) | 2,909 (15.8%) | 3,552 (23.6%) |
| Referral from another health facility or community health worker (CHW) | 24 (0.2%) | 6 (0.1%) | 184 (1.1%) | 66 (0.4%) |
| Hospitalized in the last 14 days (n, %) |  |  |  |  |
| Yes | 99 (0.7%) | 35 (0.3%) | 145 (0.8%) | 48 (0.3%) |
| No | 13,555 (99.3%) | 12,834 (99.7%) | 18,196 (99.2%) | 15,009 (99.7%) |
| Phone ownership (n, %) |  |  |  |  |
| Primary caregiver or person inside household | 12,053 (88.3%) | 11,025 (85.7%) | 14,936 (81.4%) | 12,057 (80.1%) |
| None or person outside household (including CHW) | 1,601 (11.7%) | 1,844 (14.3%) | 3,405 (18.6%) | 3,000 (19.9%) |
| **Patients (case completed electronically; per-protocol)** | **n = 8,509** | **n = 7,361** | **n = 10,331** | **n = 7,375** |
| Complaint categories (n, %) |  |  |  |  |
| Respiratory | 4,639 (54.5%) | 3,642 (49.5%) | 5,946 (57.6%) | 4,080 (55.3%) |
| Fever | 3,606 (42.4%) | 2,994 (40.7%) | 4,453 (43.1%) | 2,949 (40.0%) |
| Gastrointestinal | 2,530 (29.7%) | 2,215 (30.1%) | 2,499 (24.2%) | 2,308 (31.3%) |
| Ear nose throat mouth | 2,360 (27.7%) | 1,891 (25.7%) | 935 (9.1%) | 1,164 (15.8%) |
| Skin | 975 (11.5%) | 874 (11.9%) | 883 (8.5%) | 706 (9.6%) |
| Eye | 881 (10.4%) | 826 (11.2%) | 846 (8.2%) | 718 (9.7%) |
| Neurological | 973 (11.4%) | 646 (8.8%) | 745 (7.2%) | 636 (8.6%) |
| Others | 898 (10.6%) | 714 (9.7%) | 1039 (10.1%) | 492 (6.7%) |
| Injury musculoskeletal | 240 (2.8%) | 221 (3.0%) | 261 (2.5%) | 194 (2.6%) |

<sup>a</sup> Intervention-control analysis compares Group A Early intervention period (first five months) to Group B Control period (first five months).
<sup>b</sup> Before-after analysis compares Group B Control period (first five months) to Group B Early intervention period (second five months).
<sup>c</sup> Score (%) was calculated as the number of available commodities/services divided by the total number of commodities/services assessed per SARA methodology.
<sup>d</sup> Source: Service Availability and Readiness Assessment (SARA) conducted in December 2020.
<sup>e</sup> Source: Health Management Information System (HMIS) data for 2019-2020.
<sup>f</sup> Sum of health facilities multiplied by number of days (only Monday-Friday are considered because registration assistants did not work on weekends).

Patient enrollment was conducted over 6,556 health facility days between 1 December 2021 and 30 April 2023. Recruitment occurred on 6,105 (93%) days, with missed days primarily due to absences of registration assistants (3%) or clinicians trained on the use of the application (3%), and rarely due to IT, power and other issues (1%) (Table 1). Of 85,212 patient consultations screened, 59,921 (70%) were enrolled. Non-enrollment was attributed to ineligibility (10%); logistical barriers to consenting or enrolling patients (18%) – such as IT issues, lack of trained clinicians, adolescents coming unaccompanied to the health center, and lack of witnesses – and rarely caregiver refusal (2%) (Figure 4).

The enrolled cohort (n=59,921) was balanced between groups (A and B) in terms of sex (52% vs. 51% female) and age (65% vs. 64% aged 2-59 months) distribution. Most consultations were for new illnesses (80%), and fewer than 1% of patients had been recently hospitalized. Household access to a phone was higher in Group A (88% vs. 81%); those without a phone in the household reported access via neighbors, friends, or CHWs (Table 1).

Excluding 12,099 referral or re-attendance visits, 47,822 new consultations were eligible for the ITT analysis. Among these, the ePOCT+ algorithm was used in 70% of cases, yielding 33,576 consultations for the PP analysis of day 0 outcomes. ePOCT+ use was lower in Group B (68% in control and 64% in early intervention periods) than in Group A (75% in early and late intervention periods) (Figure 4; Figure S2, Supporting Information). Follow-up data were available for 33,238 consultations (ITT) and 23,760 consultations (PP) for day 7 outcomes. Loss to follow-up was 30% overall, and higher in Group B (34%) than in Group A (26%), consistent with phone access profile.

The most common presenting complaints were respiratory symptoms, fever and gastrointestinal issues (Table 1). The distribution of complaints was similar across study groups, although skin and ear nose mouth and throat problems were more frequently reported in Group A.

### Primary outcome: antibiotic prescription

Under routine care (Group B), antibiotics were prescribed in 70.5% of consultations for acute illness (Table 2; Fig. 5A), with highest proportions in children aged 2–59 months (73.6%) and for skin complaints (89.6%) (Fig. 5A). Use of ePOCT+ reduced antibiotic prescribing to 24.5% (Group A) and 27.5% (Group B), corresponding to absolute reductions of 46.0% (95% CI: −52.2, −39.5) in the PP intervention-control comparison and 43.0% (95% CI: −49.7, −36.4) in the before-after comparison. The largest reductions were observed in the 2-59 age group and for respiratory, ear nose throat and mouth, and skin complaints (Fig. 5B).

**Figure 5.**
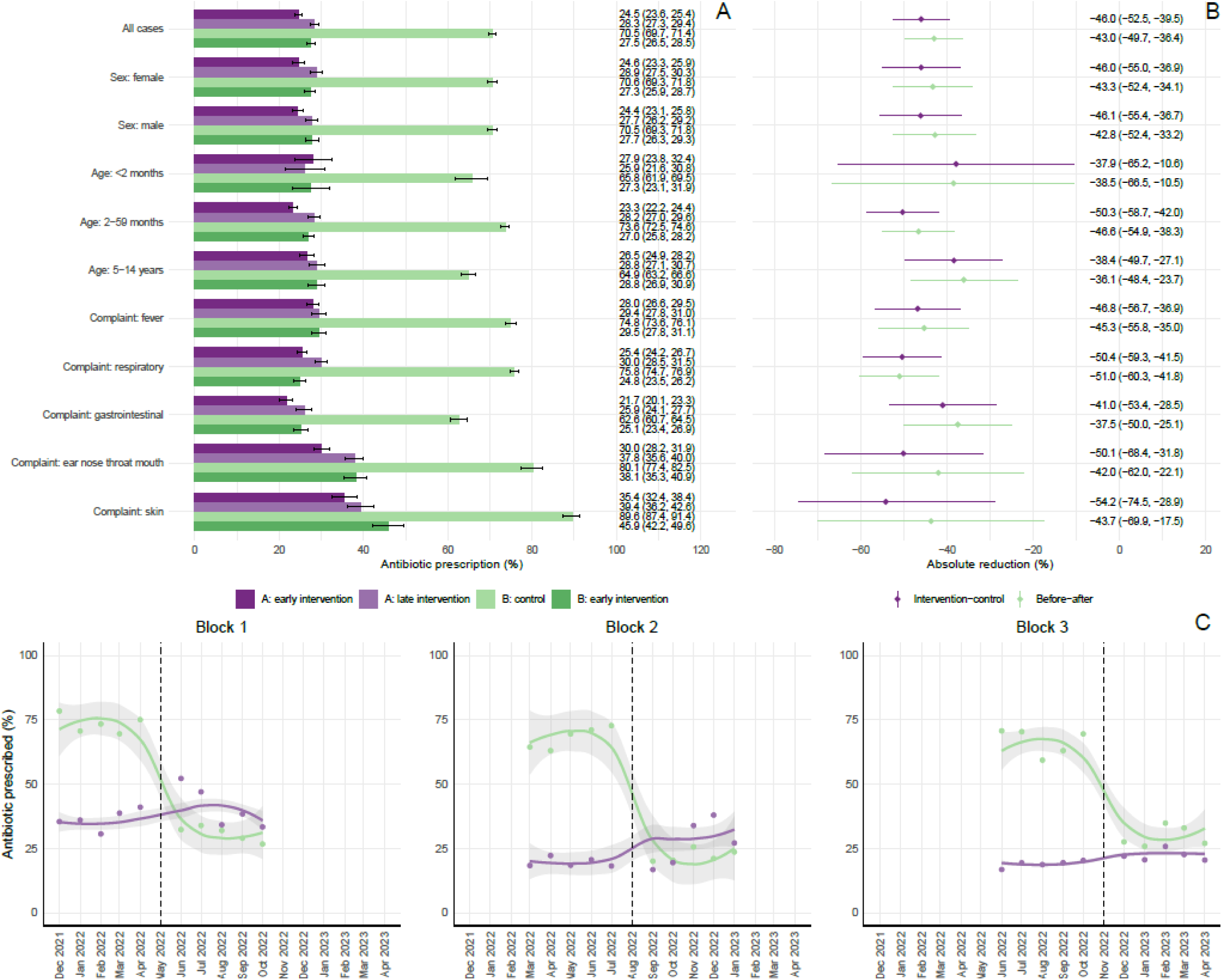
Antibiotic prescription proportions and differences across trial periods and subgroups in the per-protocol population. Panel A shows the proportion of children prescribed antibiotics at day 0 across four study periods—control (Group B), early and late intervention (Group A), and early intervention post-crossover (Group B)—overall and stratified by sex, age group, and presenting complaint. Use of ePOCT+ was associated with substantial reductions in antibiotic prescriptions in all subgroups. Panel B presents absolute reductions in antibiotic prescription between groups, comparing the early intervention phase of Group A to the control phase of Group B (intervention–control comparison), and the pre- and post-intervention phases of Group B (before–after comparison). The largest reductions were observed for skin, respiratory, and ear nose throat and mouth complaints. Panel C illustrates the monthly trends in antibiotic prescribing over time in each implementation block, highlighting drastic reduction in Group B (green) post-crossover (vertical dashed line) and consistency in sites using ePOCT+ (purple).

**Table 2.**
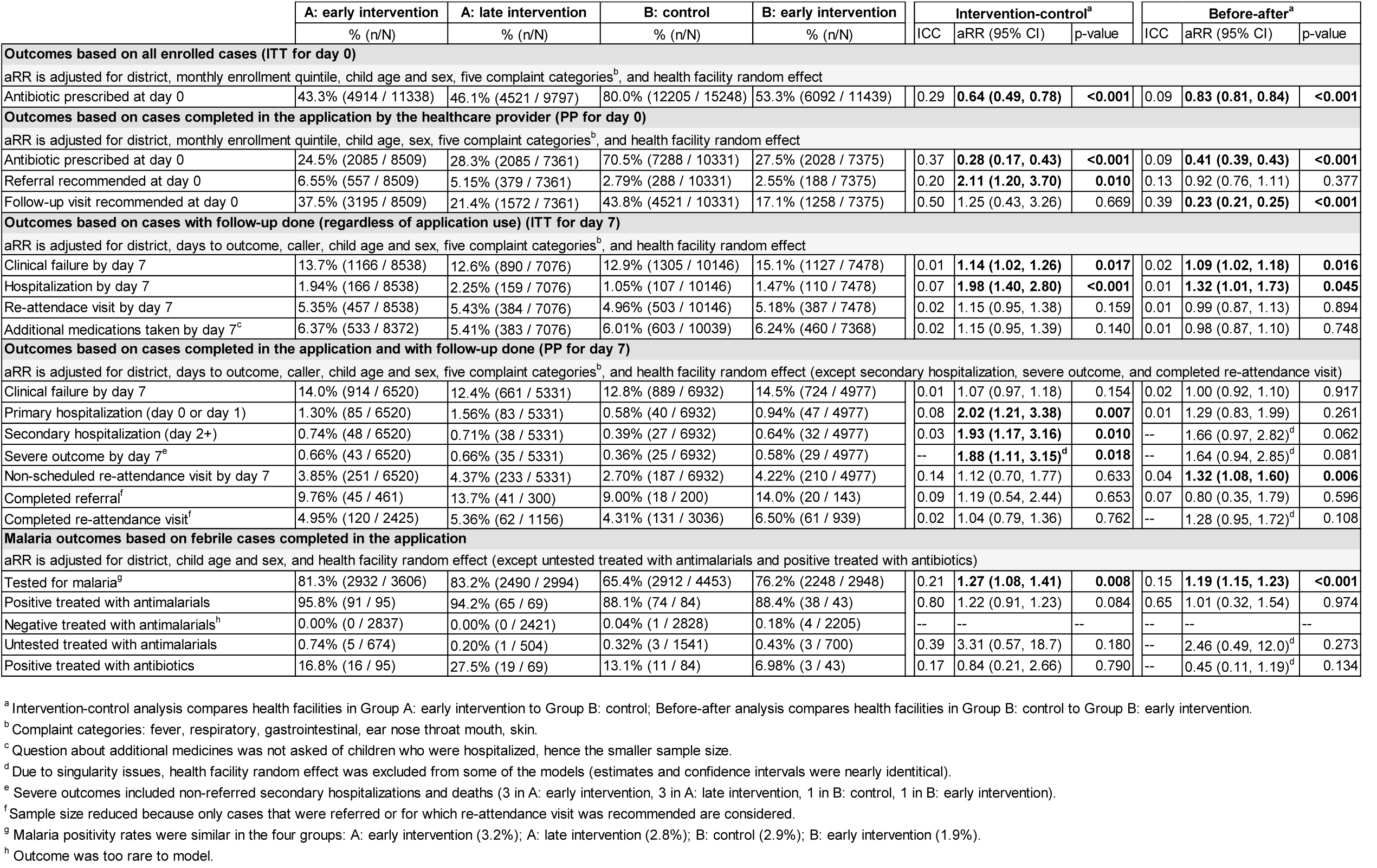
Adjusted mixed-effects logistic regression results for primary and secondary outcomes comparing ePOCT+ (intervention) to routine care (control), across the intervention– control and before–after analyses. Results are shown for intention-to-treat (ITT) and per-protocol (PP) populations at day 0 and day 7. Estimates are presented as adjusted relative risks (aRRs) with 95% confidence intervals (CIs), with variables that the model was adjusted for noted. Statistically significant results are bolded. ICC denotes intraclass correlation coefficient. Due to rare events or singularity issues, random effects were excluded from selected models. Severe outcomes comprise non-referred hospitalizations and deaths. Sample sizes vary by outcome and population. Additional footnotes provide detail on modelling specifications and subgroup definitions.]

In the ITT analysis, antibiotic prescription decreased from 80.0% to 43.3% (intervention–control) and to 53.3% (before–after), corresponding to the absolute reductions of 36.7% (95% CI: −42.1 to −31.3) and 26.7% (95% CI: −32.2 to −21.4), respectively (Table 2; Figure S3, Supporting Information). In the longitudinal analysis of Group A health facilities, antibiotic prescription remained similar throughout the 10 months of the intervention period in blocks 1 and 3; however, it increased in the second half of the intervention period in block 2 (Fig. 5C). Facility-level variation was marked, with some ePOCT+ sites maintaining high prescription proportions and some control sites demonstrating low baseline antibiotic use (Figure S4, Supporting Information).

### Secondary outcomes: clinical failure, referral, hospitalization, malaria management, additional medications

In the adjusted PP multivariable analysis, clinical failure did not differ significantly among children managed according to routine care (12.8%) vs. with ePOCT+ (aRR 1.07; 95% CI 0.97– 1.18 [intervention–control]; aRR 1.00; 95% CI 0.92–1.10 [before–after]). In the ITT analysis, effect estimates were modestly higher in both comparisons, but within the non-inferiority margins (aRR 1.14; 95% CI 1.02–1.26 and 1.09; 95% CI 1.02–1.18, respectively) (Table 2). No major differences were observed across subgroups, though failure rates in the ePOCT+ group were slightly higher for skin conditions, which typically take longer to resolve (Figure S5, Supporting Information).

In the intervention-control comparison, referral rates were two times lower in the control arm (2.8%) compared to the intervention arm (2.8% vs. 6.6%; aRR 2.11; 95% CI 1.20–3.70) (Table 2), likely due to better detection of severe cases (Figure 6A). Primary and secondary hospitalizations (0.6% and 0.4%, respectively at baseline) were also approximately two times in the intervention arm (aRR 2.02; 95% CI 1.21–3.38 and aRR 1.93; 95% CI 1.17–3.16, respectively). However, the adherence to referral advice was low (<15%). Severe outcomes, comprised primarily of non-referred secondary hospitalizations (with very few deaths), were also more frequent in the intervention group (0.66% vs. 0.36%; aRR 1.87; 95% CI 1.11–3.15). No differences in referrals, hospitalizations or severe outcomes were observed in the before-after comparison (Table 2). Most of the children with non-referred secondary hospitalization had been cured or improved by day 7. Among these children, clinical outcomes were similar regardless of antibiotic treatment at day 0 (Table S1, Supporting Information).

**Figure 6.**
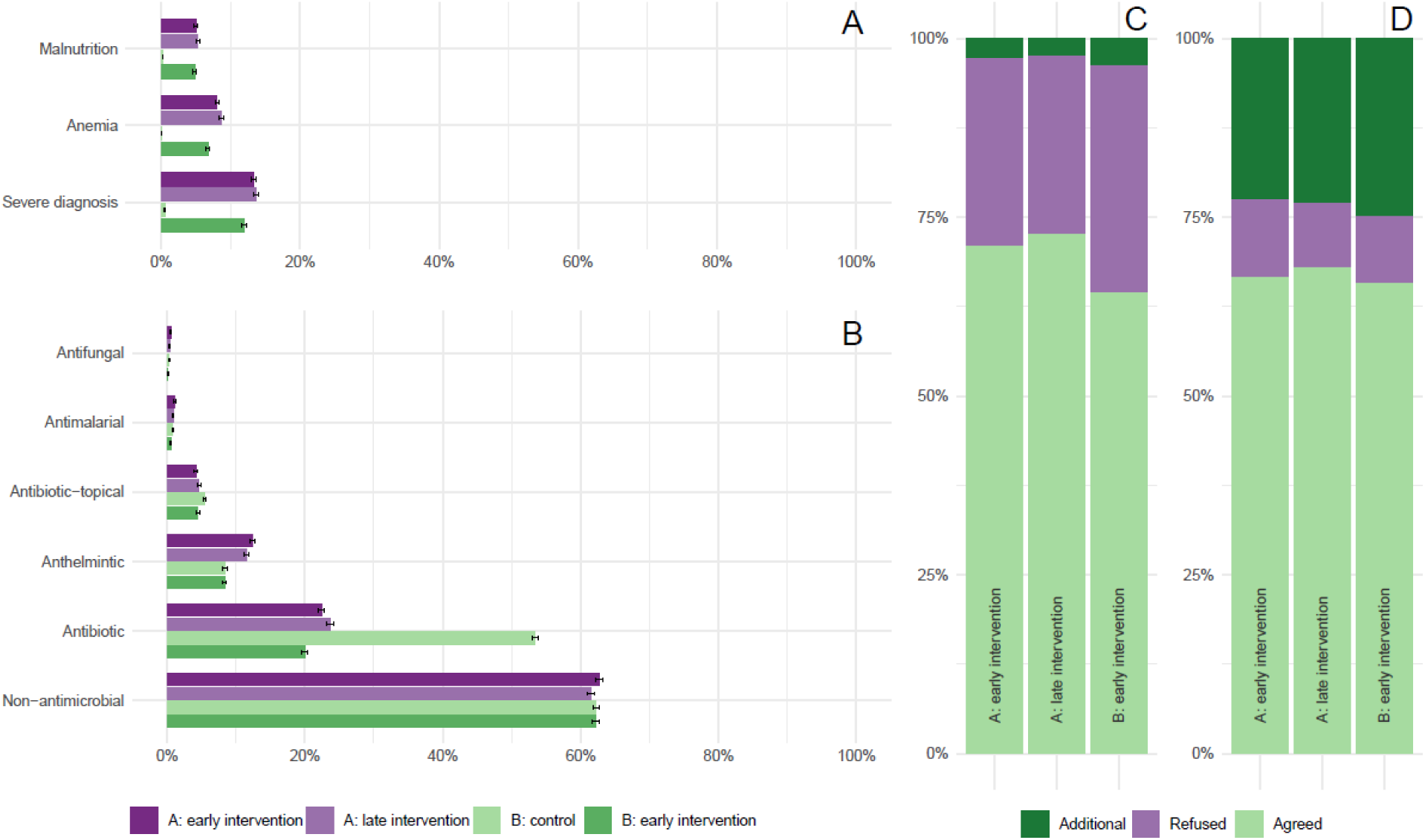
Impact of ePOCT+ on diagnosis, treatment, and adherence to algorithm recommendations. Panel A shows the proportion of consultations in which severe diagnoses, anemia, and malnutrition were identified. Use of ePOCT+ led to marked increases in the detection of all three conditions compared to routine care. Panel B presents the distribution of medication prescriptions by category, including antibiotics, antimalarials, antifungals, and non-antimicrobial agents. Aside from a reduction in antibiotic use, the overall prescription pattern remained stable across groups. Panel C displays clinician agreement with algorithm-suggested diagnoses. While most were accepted, approximately one-quarter were rejected across intervention periods. Panel D shows clinician agreement with algorithm-suggested antibiotics. Rejection of proposed antibiotics was rare, but around 25% of prescribed antibiotics were added manually, indicating partial algorithm adherence.

Malaria testing of febrile cases improved with the use of ePOCT+ from a baseline of 65.4% in the control group (aRR 1.27; 95% CI 1.08–1.41), though treatment adherence was already high (>88%) and did not change significantly. A very small percentage of children (<6.5%) subsequently took medications in addition to those prescribed to them at the study consultation, with no difference between the ePOCT+ and routine care arms (Table 2). Results of unadjusted analyses are provided in Table S2, Supporting Information.

### Impact of ePOCT+ on diagnosis and treatment decisions

Routine use of anthropometric and hemoglobin measurements with ePOCT+ improved detection of malnutrition (from <0.5% to ∼5%) and anemia (from 0% to ∼8%). Severe diagnoses were also identified more frequently (<1% vs. 12-13%) (Figure 6A). Apart from the reduction in oral and parenteral antibiotics, there were no significant changes in medicine prescriptions (Figure 6B). Some gaps were noted in compliance with ePOCT+ recommendations. Clinicians rejected diagnoses proposed by the algorithm approximately 25% of the time, while manually added diagnoses were rare (Figure 6C). On the other hand, rejection of proposed antibiotics was rare, but manual addition when not proposed by ePOCT+ constituted approximately 25% of all prescribed antibiotics (Figure 6D).

## Discussion

This pragmatic trial demonstrated a substantial reduction in antibiotic prescription rates for children aged under 15 years managed in primary care settings using a CDSA integrated with point-of-care tests and supported by clinical mentorship. In the per-protocol analysis, antibiotic prescriptions declined nearly three-fold; in the intention-to-treat analysis, the reduction was two-fold. The most pronounced reductions were observed in children aged 2–59 months— representing over 65% of the outpatient pediatric caseload—and among those presenting with respiratory, skin, or ear nose throat and mouth complaints, which together accounted for the majority of consultations. These findings are consistent with the cluster-randomized trial of ePOCT+ in Tanzania, where antibiotic prescribing also declined approximately three-fold from 70% at baseline in the PP analysis and two-fold in the ITT analysis [24]. However, in a recent pre-post study in Kenya and Senegal of a similar algorithm limited to the under 5 age range, a more modest reduction in antibiotic prescription (15-23%) was observed. Notably, the algorithm used in that study included pulse oximetry but excluded hemoglobin and CRP diagnostic tests, and the implementation and clinical mentorship strategies differed [42]. These factors possibly contributed to improved antibiotic stewardship in the present study.

The immediate effect of the intervention appeared to be sustained across a ten-month longitudinal assessment. In our study, antibiotic reductions were not accompanied by increased prescribing of other medications, suggesting that clinicians did not feel the need to compensate by substituting other medications. Similarly, parents did not give additional medicines to their children at home, except in a minority of cases, balanced across study arms.

Algorithm adherence, however, remained suboptimal. Nearly one-third of the prescribed antibiotics were added manually by clinicians and not recommended by the algorithm. This high rate suggests that this was not related to special clinical cases outside guidelines but rather persistent reliance on clinical judgement and the need for ongoing mentorship. Antibiotic prescribing rates—and the rate of manual additions when not proposed by ePOCT+—varied widely across facilities and individual clinicians. These findings suggest that with stronger supervision and better adherence, prescribing rates could fall even further, approaching the 11% observed in prior efficacy studies in controlled research conditions of an earlier ePOCT version [22]. Further analyses include exploring the relative contributions of the algorithm, CRP test, and monitoring and clinical mentorship enabled by the real-time dashboards on antibiotic prescriptions.

Importantly, ePOCT+ improved the detection of malnutrition and anemia through routine assessment of anthropometrics and hemoglobin. Malaria testing rates also increased significantly, although malaria treatment adherence was already high and remained unchanged. These improvements reflect how algorithm-guided care may contribute to overall diagnostic quality.

Despite the impact demonstrated in the per-protocol population, it is important to note that 25– 30% of the enrolled cases were not managed with ePOCT+. Lack of integration among reporting tools, with paper-based tools (IMCI registers, health insurance forms) and, in some health centers, electronic medical records, being used in parallel, made workflows time-consuming. Staffing constraints varied widely, with the most pronounced challenges in remote Nyamasheke district (group B), where high staff turnover and patient volume hampered consistent use of the application. These systemic factors underscore the importance of harmonizing clinical, administrative, and reporting tools into a unified digital system, particularly in low-resource settings. Individual-level factors, including digital literacy and motivation of clinicians to use CDSAs, warrant further investigation.

Although the findings support clinical safety of ePOCT+, the study was not powered to evaluate these secondary outcomes. Clinical cure rates were similar across groups in both intervention-control and before-after comparisons. Non-inferiority in clinical outcomes of the ePOCT+ algorithm was also confirmed in the cluster-randomized trial in Tanzania [24]. While referrals, hospitalizations, and severe outcomes did not differ in the before–after comparison, they were more frequent in the ePOCT+ group in the intervention–control comparison. This most likely reflects an imbalance in baseline severity between groups, as suggested by higher rates of severe diagnoses in Group A. Despite the inclusion of oxygen saturation and hemoglobin measurements in ePOCT+ to improve detection of severe illnesses, referrals did not increase in the Tanzanian trial [24], nor in the Kenyan and Senegalese study of a similar algorithm for under five children with pulse oximetry [42]. Complementary concerning findings were low referral completion among caregivers of children and frequent non-referred hospitalizations, indicating remaining challenges both on behalf of clinicians in identifying severe cases and on behalf of patients reaching the hospital when referred. These patterns warrant further study.

Our study has several strengths and limitations. Strengths include a large sample size, multiple comparisons, and broad patient inclusion criteria, making the findings generalizable to the general pediatric outpatient population. Limitations include a clinician self-reported measure of antibiotic prescription, an approach often used in pragmatic studies [43], and relatively high loss-to-follow up (∼30%); although not dissimilar to that of other SSA studies [44]. The clinician self-reported nature of the antibiotic prescription outcome may mean that the reduction we found in the PP analysis is over-estimated, and most likely the real reduction is closer to that found in the ITT analysis, as was the case in the ePOCT+ study in Tanzania [21].

Although it could be perceived as a limitation, the use of a non-randomized, staggered implementation design was a deliberate methodological decision aligned with the study’s pragmatic objectives. Rather than prioritizing experimental control, the design aimed to replicate how the Rwandan Ministry of Health would likely scale the intervention in routine primary care settings. This real-world approach enhances the study’s external validity and supports its relevance for health policy and systems strengthening efforts [45]. This design choice is consistent with the PRECIS-2 framework, which advocates for aligning trial characteristics with the intended purpose of the intervention—particularly when the goal is to inform large-scale implementation or policy decisions [46]. While randomized controlled trials remain the gold standard for determining efficacy, non-randomized designs are increasingly recognized as appropriate for evaluating complex interventions’ performance under routine operational conditions, with a focus on implementation fidelity, acceptability, scalability, and health system integration [47,48]. This sequential approach reflects the widely endorsed explanatory– pragmatic continuum, which aims to bridge the gap between innovation and policy uptake in global health research [46,49]. By adopting this perspective, the study contributes not only to understanding the intervention’s effectiveness in a new setting but also to broader discussions about the sustainable integration of digital health tools into public sector health systems.

Although the absence of randomization introduces some limitations in terms of internal validity, the study design enhances generalizability and aligns with internationally recognized guidance for evaluating complex, context-dependent interventions [47,48].

## Conclusion

In this pragmatic trial, the implementation of ePOCT+ substantially reduced antibiotic prescribing in pediatric outpatient care in Rwanda, without compromising clinical recovery. If scaled more broadly, ePOCT+ has the potential to strengthen adherence to clinical guidelines and support national efforts to promote antibiotic stewardship and combat antimicrobial resistance. Tackling antimicrobial resistance is a strategic priority for the Rwandan health sector, as outlined in the national action plan on AMR [30], the updated national standard treatment guidelines [31], and the WHO AWaRe classification [50]—all of which emphasize antibiotic stewardship and rational prescribing. In this context, integrating ePOCT+ and other decision support tools into Rwanda’s national electronic medical record platform in accordance with the national digital health strategy [32], and ensuring that all clinical and reporting functions can be performed through a unified system, represents a critical next step. Such integration could streamline service delivery, improve data quality, and promote more consistent, evidence-based care at scale.

## Data Availability

De-identified data used in the analysis can be found at https://doi.org/10.5281/zenodo.15870974. Note: There is an embargo period until the end of the year to allow for publication of the article.

https://zenodo.org/records/15870974

## Acknowledgments

We thank the study patients and caregivers for participating in the study and the nurses, health facility administrators and district staff for facilitating the use of ePOCT+ and the medAL-*reader* application in the health centers. We also thank Antoinette Makuza, Jonathan Niyonzima, Gilbert Rukundo, Serge Zimulinda, Aicha Yusuf, Alex Rosen, Theophile Dusengumuremyi, Clementine Tumukunde, Francine Bayisenge, and Roberta Minotti for their early contributions to setting up the study and accompanying the nurses. We appreciate members of the steering committee, Rwanda Biomedical Center, the Ministry of Health, and Rwanda Pediatric Association for their continued guidance throughout the project. Special thanks to Sabin Nsanzimana, Muhammed Semakula and Claude Muvunyi for their support. We also thank the registration assistants who recruited patients in the health centers and research staff who conducted the day 7 phone calls. We also thank members of the Unisante IT team (Sylvain Schaufelberger, Gregory Martin, and Julien Thabard) and Wavemind (Alain Fresco, Emmanuel Barchichat, and Quentin Girard). Additional thanks to Kristina Keitel, Mary-Anne Hartley, Fenella Beynon, Lisa Cleveley and members of the DYNAMIC Tanzania team (Alix Miauton, Chacha Mangu, Sabine Renggli, Lameck Luwanda, Godfrey Kavishe, Geofrey Ashery, Margaret Jorram, Peter Agrea, Ibrahim Mtebene).

## Author Contributions

AVK, VDA, and KW were responsible for the study design. AVK, RT and VDA developed the statistical analysis plan. AVK and VF were responsible for data curation. AVK carried out the formal analysis. VDA, KW and AVK acquired the funding. AVK, VPR, JH, MN, LC, EK, CH, AI, GAL, VF carried out the investigation and collected the data. AVK, MN, LC, VF and AV were responsible for project administration. AVK, VDA, and JCSN supervised the project. AVK wrote the original draft of the manuscript. VPR, JH, MN, LC, EK, CH, AI, GAL, VF, AV, MALP, KW, LT, JCSN, and VDA reviewed and edited subsequent drafts of the manuscript. AVK had full access to all of the data in the study and takes responsibility for the integrity of the data and the accuracy of the data analysis.

